# SARS-CoV-2 Seroprevalence in 12 Cities of India from July-December 2020

**DOI:** 10.1101/2021.03.19.21253429

**Authors:** Arokiaswamy Velumani, Chaitali Nikam, Wilson Suraweera, Sze Hang Fu, Hellen Gelband, Patrick Brown, Isaac Bogoch, Nico Nagelkerke, Prabhat Jha

## Abstract

**Objectives:** We sought to understand the spread of SARS-CoV-2 infection in urban India, which has surprisingly low COVID-19 deaths.

**Design:** Cross-sectional and trend analyses of seroprevalence in self-referred test populations, and of reported cases and COVID mortality data.

**Participants:** 448,518 self-referred individuals using a nationwide chain of private laboratories with central testing of SARS-CoV-2 antibodies and publicly available case and mortality data.

**Setting:** 12 populous cities with nearly 92 million total population.

**Main outcome measures:** Seropositivity trends and predictors (using a Bayesian geospatial model) and prevalence derived from mortality data and infection fatality rates (IFR).

**Results:** For the whole of India, 31% of the self-referred individuals undergoing antibody testing were seropositive for SARS-CoV-2 antibodies. Seropositivity was higher in females (35%) than in males (30%) overall and in nearly every age group. In these 12 cities, seroprevalence rose from about 18% in July to 41% by December, with steeper increases at ages <20 and 20-44 years than at older ages. The “M-shaped” age pattern is consistent with intergenerational transmission. Areas of higher childhood measles vaccination in earlier years had lower seropositivity. The patterns of increase in seropositivity and in peak cases and deaths varied substantially across cities. In Delhi, death rates and cases first peaked in June and again in November; Chennai had a single peak in July. Based local IFRs and COVID deaths (adjusted for undercounts), we estimate that 43%-65% of adults above age 20 had been infected (range of mid-estimates of 12%-77%) corresponding 26 to 36 million infected adults in these cities, or an average of 9-12 infected adults per confirmed case.

**Conclusion:** Even with relatively low death rates, the large cities of India had remarkably high levels of SARS-CoV-2 infection. Vaccination strategies need to consider widespread intergenerational transmission.

## Introduction

As of early March 2021, India had over 11 million confirmed SARS-CoV-2 infections (COVID cases), second only to the United States (US), and nearly 160,000 reported COVID deaths. Cases and deaths in India constitute about one in nine and one in thirteen, respectively, of worldwide totals.^1^ India’s COVID mortality rate adjusted for population size and age structure is substantially lower than that in the US or Europe.^2^ Within India, the reported cases have been concentrated in cities, with New Delhi, Bengaluru, Pune, Mumbai, Thane, and Chennai together reporting more than two million individuals with positive PCR tests.^3^ Reported COVID deaths are similarly concentrated, although both cases and deaths in rural areas are likely under-reported due to lower access to viral testing and lower levels of registration and certification of deaths than in urban areas. Most rural deaths occur at home without medical attention at the time of death.^4^

The transmission patterns of SARS-CoV-2 in India are likely to differ substantially from high-income countries or even from China,^5^ given that multiple generations typically live together. Moreover, high population density, particularly in urban areas, complicates physical distancing both at home and outside.

Serological surveys in focal areas of India report SARS-CoV-2 antibody seropositivity of 20-50% of adults.^6-9^ However, standardized seroprevalence estimates across the many diverse settings of urban India have not yet been documented. Here, we report results of serological testing from nearly 450,000 self-referred patients using a nationwide chain of private laboratories with central testing of IgG antibodies. We combine these data with recorded case counts and use demographic analyses of baseline and COVID-19 mortality to estimate mortality-derived infection rates in 12 of the most populous cities that have reasonably complete recording of all deaths. We also examine, on an ecological level, the association of past childhood vaccination^10^ and other risk factors^11^ to adult SARS-CoV-2 infection.

## Methods

### Population tested

We included 448,518 people of all ages who underwent testing for SARS-CoV-2 antibodies between June 12, 2020 and December 31, 2020 by Thyrocare Laboratories.^12^ Thyrocare conducts central laboratory testing in Navi Mumbai from over 2200 franchised collection centers in all major cities in India. Thyrocare charges 600-750 Rupees (US$8-10) for an antibody test. The available data included the type of assay used, sample collection date, age, sex, and area postal pincode, but no personal identifiers. Motives for testing were not ascertained. Based on Indian income distribution in urban areas,^13^ it is likely that only the higher socioeconomic strata (SES) can afford these costs.

### SARS-CoV-2 IgG serological assays

The SARS-CoV-2 antibody assays involved collection of 3 ml of serum, shipment overnight to Navi Mumbai, and central high-throughput analyses using one or both of two widely-used assays that detect antibodies to the SARS-Cov-2 nucleocapsid protein: the Abbott SARS-CoV-2 chemiluminescent microparticle immunoassay for IgG (positive cutoff: >=1.40),^14^ and the Cobas, Roche Elecsys Anti-SARS-CoV-2 electrochemiluminescence immunoassay for total immunoglobulin (most of which is IgG; positive cutoff: >=1.0).^15^ For both assays, the laboratory followed the manufacturers’ quality control procedures.^12^

### SARS-CoV-2 PCR and rapid antigen testing, and mortality in 12 metropolitan areas

We compared seropositivity from the laboratory results to cases and deaths in 12 cities with reasonably high completeness (>90%) of reporting deaths in pre-pandemic years, as defined by the Registrar General of India (RGI).^16^ These cities have a collective population nearly 92 million (61 million adults above age 20 years), including the megacities of Mumbai, Delhi, and Kolkata, as well as Chennai, Bangalore, Ahmedabad, Pune, Surat, Jaipur, Nagpur, Coimbatore, and Visakhapatnam. For all these cities, we obtained SARS-CoV-2 polymerase chain reaction (PCR) or rapid antigen testing results (some states used rapid tests in June and then switched to PCR testing). We do not distinguish between the two methods. COVID death reporting follows World Health Organization classification guidelines.^17^ Both cases and death data were from the COVID19India.org,^3^ a volunteer group that collects data daily from all official data sources. We compared these data with daily official statistics globally,^1^ or published by the Government of India,^18^ filling gaps through web searches of local bulletins and social media. We derived population and death rates from vital statistics.^16^

### Data Analysis

The main calculations are of age- and sex-specific seropositivity nationally, and confirmed cases and COVID deaths per population over the age of 20 years, of the assayed participants. About 40% of the population tested by Thyrocare were in the 12 included cities. Because the tested population was not representative, we conducted a mortality-based estimate to calculate the prevalence of infection in each city. The relation of the age-specific SARS-CoV-2 prevalence (P) to COVID attributable mortality is P=DR/IFR where DR is the COVID death rates and IFR is the assumed infection fatality rate. The IFRs used values of 0.29%, 0.263% and 0.21%, drawn, respectively, from detailed serosurveys in Bangalore, Mumbai, and Pune.^6-8^ We applied a reasonably large 30% correction factor based on RGI data on death reporting completeness, and previous analyses of under-reporting in selected low and middle-income countries^2,19^ to arrive at adjusted COVID deaths which we then used to calculate the infection prevalence. Sensitivity analyses examined 0 to 50% undercount scenarios.

We linked the 1415 pincodes (83% of the total) from the serology data to a commercial pincode geocoding dataset,^20^ representing a wide geographic distribution across India (363 pincodes for the 12 cities, and 1054 for all other areas, supplementary figure 4), and to other spatial datasets. We used a geostatistical Bayesian logistic regression model to estimate the spatially smoothed probability of seropositivity, with the seropositive cases (based on positive results for either antibody assay) as the outcome and the total tested as the denominator for each pincode (yielding odds ratios [OR]). The model examined migrants from another state, population less than 20 years old, mean of particulate matter of 2.5 micron size (PM_2.5_; drawn from 2015-17 satellite-based exposure data^21^), and nationally representative estimates of childhood polio and measles vaccination from the National Family Health Survey 4 conducted in 2015-16.^10^ Because childhood vaccination was correlated, we examined polio and measles separately. The model also adjusted for sex, non-linear effect of age, percentage of urban settlement, population size, population density in the pincode, subdistrict-level solid-fuel use (as a measure of rural residence and lower socioeconomic status), female illiteracy, and language (as a proxy for geographic regions). We derived co-variates mostly from 2011 Indian census or WorldPop gridded data.^22^ To reduce the “noise” in the resulting geographic estimates, we added smoothly varying spatial random effects (i.e., spatial autocorrelation or clustering) and independent unit-level random effects.^23^ Additional details of the analyses are provided in the supplementary appendix.

## Patient and public involvement

Owing to the nature of this research, no patients or members of the public were involved in the design or reporting of this study.

## Results

For the whole of India, 31% (140,631) of the 448,518 self-referred people who tested were seropositive for SARS-CoV-2 antibodies on either assay. Seropositivity to SARS-CoV-2 rose from about 16% in July until December, by which time nearly 50% of the tested population was seropositive. Of those tested, 218, 111 used the Abbott assay, 103,221 used the Roche assay, and 127,186 used both. We present results for either assay interchangeably as the concordance of the two assays was high: arbitrarily choosing the more widely used Abbot assay as the reference, the Roche assay showed a sensitivity and specificity of 87% and 97%, respectively (Kappa agreement 0.879; 95%CI 0.876, 0.882). Analyses using dual positive individuals yielded similar results (data not shown). The standardized titers of the two assays yielded comparable age-specific results, with wider divergence at older ages (data not shown).

Seropositivity from July 1 to December 31, 2020 was higher in females (35%) than in males (30%) in nearly every age group (Figure 1 panel A, supplementary table 1). The “M-shaped” age pattern is broadly consistent with widespread intergenerational transmission: the sex-specific OR for seropositivity, using age 20 as the reference age and adjusted for location and various other covariates, rose in children to age 14, fell by age 20, and then rose to peaks at age 60 in females and 70 in males after which it declined (panel B). The seropositivity rates rose from July to December for each of the age groups <20, 20-44, 45-69 and 70+ (panel C), with the fastest growth at ages <20 and 20-44 years. Over 61% of the tested population were aged 20-44 years (supplementary table 1). Logistic regression of the daily new seropositive cases showed fluctuations nationally in seropositivity, with the peak of daily cases broadly synchronous with a September peak of PCR confirmed cases and deaths, followed by declining daily counts (panel D).

**Figure 1.**
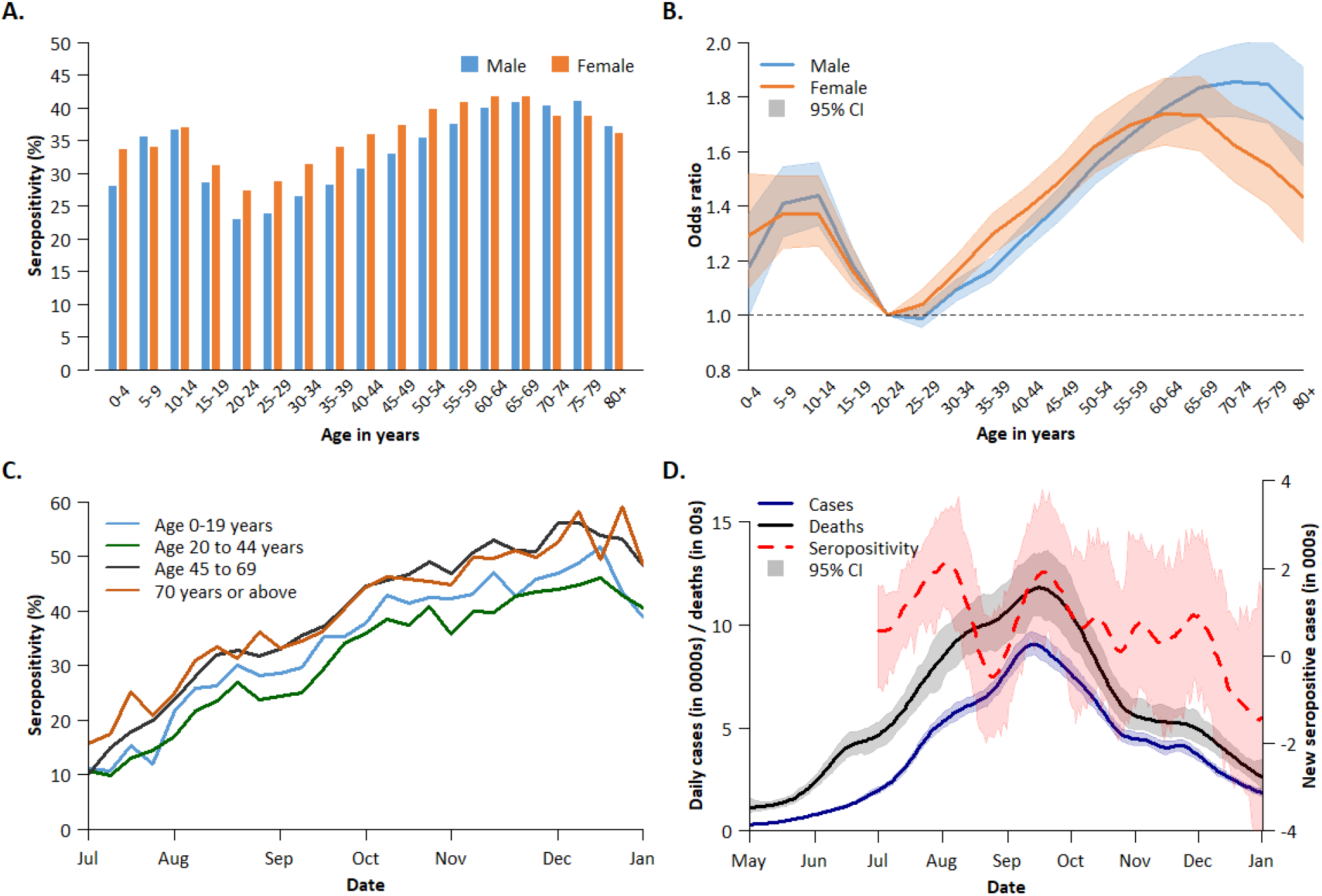
Seropositivity (%) by sex and age from July to December,2020; ORs of infection by sex and by time, and new seropositive cases and COVID cases and deaths by time. **A.** Age and sex-specific seropositivity. **B**. ORs of seropositivity by sex. **C**. Trends over time by age group, both sexes combined. **D**. Daily incidence of seropositivity, COVID cases, and deaths over time and uncertainty intervals. In Panel B, the OR uses age 20 as the reference in logistic regressions. Panel D explores temporal effects using a non-spatial logistic regression to model OR of seropositive prevalence in reference to July 1^st^ and non-spatial Poisson regressions to model the RRs of COVID cases and deaths in reference to May 1^st^, using second order random walks to model the non-linear temporal effects. The ORs were converted to absolute seropositive incidence (daily difference of seropositive prevalence) using the total number of serology tests, and RRs were converted to absolute COVID cases and deaths using the total Indian population in 2020 (see supplementary appendix).

By December, seropositivity was 41% in the 12 cities but with marked variation in monthly growth from July across the cities (supplementary table 1 and supplementary figures 1 and 2). There was a steady increase in seropositivity in all cities, except in Surat, Vishakhapatnam, and Nagpur, which showed earlier peaks in August, September, and October, respectively. Daily changes in seropositive case were similar (supplementary figure 3). Table 1 presents the cumulative totals of PCR/rapid test confirmed cases and COVID deaths in the 12 cities through December 31, 2020. The 12 cities comprised just under a third of national totals of COVID cases, and over 40% of all COVID deaths. The highest COVID death rate per 100,000 population was in Pune (82) and Nagpur (69) and the lowest was in Vishakhapatnam (13) and Jaipur (8).

**Table 1.** Seropositivity to SARS-CoV-2, PCR confirmed cases and COVID deaths in 12 large cities in India.

| City (state, population in millions) | Number tested for antibody | Age adjusted seropositivity (%) * |  |  |  |  |  |  | Mortality-derived prevalence † |  |  | PCR/rapid test confirmed cases and COVID deaths |  |  |  |
| --- | --- | --- | --- | --- | --- | --- | --- | --- | --- | --- | --- | --- | --- | --- | --- |
|  |  | Jul | Aug | Sep | Oct | Nov | Dec | Avg monthly growth | High IFR (0.29%) | Mid IFR (0.26%) | Low IFR (0.21%) | Cases (000s) | Deaths | Case rate/ 1000 Pop | Death rate/ 100,000 Pop |
| **Mumbai (MH,18.4)** | 38,581 | 28.9 | 31.2 | 34.4 | 41.3 | 38.9 | 36.8 | 4.0 | 55.7 | 61.5 | 77.0 | 670 | 19,091 | 18.0 | 51.3 |
| **Delhi (DH,16.3)** | 25,975 | 25.2 | 22.8 | 27.8 | 34.3 | 43.7 | 54.9 | 13.0 | 43.8 | 48.3 | 60.5 | 625 | 10,536 | 32.8 | 55.2 |
| **Kolkata (WB,14.1)** | 35,013 | 18.6 | 25.5 | 28.5 | 36.5 | 54.6 | 50.0 | 16.5 | 55.6 | 61.3 | 76.8 | 276 | 5,960 | 12.2 | 26.3 |
| **Chennai (TN,8.7)** | 12,528 | 23.6 | 19.5 | 28.3 | 34.5 | 36.2 | 34.9 | 6.5 | 42.6 | 47.0 | 58.8 | 296 | 5,118 | 24.0 | 41.4 |
| **Bangalore (KN,8.5)** | 25,406 | 14.0 | 20.4 | 28.1 | 33.4 | 38.0 | 42.3 | 18.4 | 30.5 | 33.6 | 42.1 | 425 | 4,621 | 29.8 | 32.4 |
| **Ahmedabad (GJ,6.4)** | 7,986 | 11.1 | 23.4 | 26.9 | 31.2 | 36.6 | 48.4 | 24.6 | 26.1 | 28.8 | 36.1 | 67 | 2,366 | 7.8 | 27.5 |
| **Pune (MH,5.1)** | 9,794 | 12.8 | 22.1 | 31.4 | 34.5 | 30.7 | 24.4 | 10.7 | 69.9 | 77.1 | 96.5 | 373 | 7,767 | 39.5 | 82.4 |
| **Surat (GJ,4.6)** | 10,058 | 32.0 | 47.9 | 35.5 | 38.8 | 33.2 | 32.8 | 0.4 | 13.7 | 15.2 | 19.0 | 49 | 966 | 8.1 | 15.9 |
| **Jaipur (RJ,3.1)** | 4,375 | 13.0 | 16.7 | 22.9 | 44.4 | 50.9 | 63.8 | 26.5 | 10.6 | 11.7 | 14.7 | 57 | 501 | 8.6 | 7.6 |
| **Nagpur (MH,2.5)** | 3,785 | 10.1 | 10.5 | 32.1 | 49.7 | 38.5 | 41.8 | 23.7 | 64.8 | 71.5 | 89.5 | 126 | 3,204 | 27.0 | 68.9 |
| **Coimbatore (TN,2.2)** | 7,892 | 8.2 | 8.2 | 19.0 | 34.1 | 38.2 | 32.8 | 23.1 | 15.5 | 17.0 | 21.3 | 52 | 653 | 15.1 | 18.9 |
| **Visakhapatnam (AP,1.7)** | 3,878 | 8.73 | 37.3 | 49.8 | 51.5 | 36.9 | 33.8 | 22.6 | 17.5 | 19.3 | 24.2 | 59 | 551 | 13.8 | 12.8 |
| **Three mega cities (48.8)** | **99,563** | **24.9** | **26.6** | **31.0** | **38.4** | **46.1** | **47.2** | **10.6** | 52.2 | 57.5 | 72.0 | **1,572** | **35,587** | 19.9 | 45.1 |
| **Total 12 cities (91.5)** | **185,271** | 17.8 | 23.8 | 30.4 | 38.7 | 39.7 | 41.4 | 14.1 | 43.1 | 47.5 | 59.5 | **3,077** | **61,334** | 20.7 | 41.2 |
| **All other areas** | **263,247** | 11.7 | 25.1 | 32.9 | 44.6 | 47.6 | 51.2 | 24.7 | 24.9 | 27.5 | 34.4 | **7,248** | **87,684** | 6.0 | 7.3 |
| **All of India** | **448,518** | 18.1 | 26.4 | 31.6 | 39.9 | 44.4 | 46.5 | 15.7 | 31.5 | 34.8 | 43.5 | **10,093** | **149,018** | 7.6 | 11.0 |

**Notes:**
\* Seropositivity rates of cities were adjusted to overall age distribution of all tested population from July-December 2020. We exclude the 1816 tested in June. Cities are listed in order of the population size. Ahmedabad metropolitan area includes Ahmedabad and Gandhi Nagar districts. Bangalore includes Tumakuru, Bangalore, Bengaluru Rural. Chennai includes Chennai, Tiruvallur and Kanchipuram. Kolkata includes Kolkata, North 24 Parganas, South 24 Parganas districts. Mumbai Includes Mumbai city, Mumbai Suburban, Palghar, Raigarh and Thane. Abbreviations: IFR Infection fatality rate; MH Maharashtra; WB West Bengal; TN Tamil Nadu; KN Karnataka; GJ Gujarat; RJ Rajasthan; BH Bihar; AP Andhra Pradesh. † Refer to Supplementary Appendix for details used to calculate the low and high IFR.

The overall patterns for all of India revealed a mid-September peak for cases and deaths (Figure 1 panel D), but with marked spatial variation (supplementary figure 5). Figure 2 illustrates this for Delhi, Pune, and Chennai. Delhi had two peaks of death rates and case rates, one in June and another in November, with seroprevalence in the self-referred population rising slowly to December. Pune’s case rate peaked in September, after deaths had peaked in August with seroprevalence in the self-referred population peaking in October. Chennai’s death and case rates peaked two months earlier in July with a continuous increase in seroprevalence in the self-referred population. The Pearson correlations were strong between city-specific COVID deaths and cases (0.84, p <.0001) and by week within each of the cities (data not shown) but only moderately between seropositivity and deaths (0.45, p <.0001).

**Figure 2.**
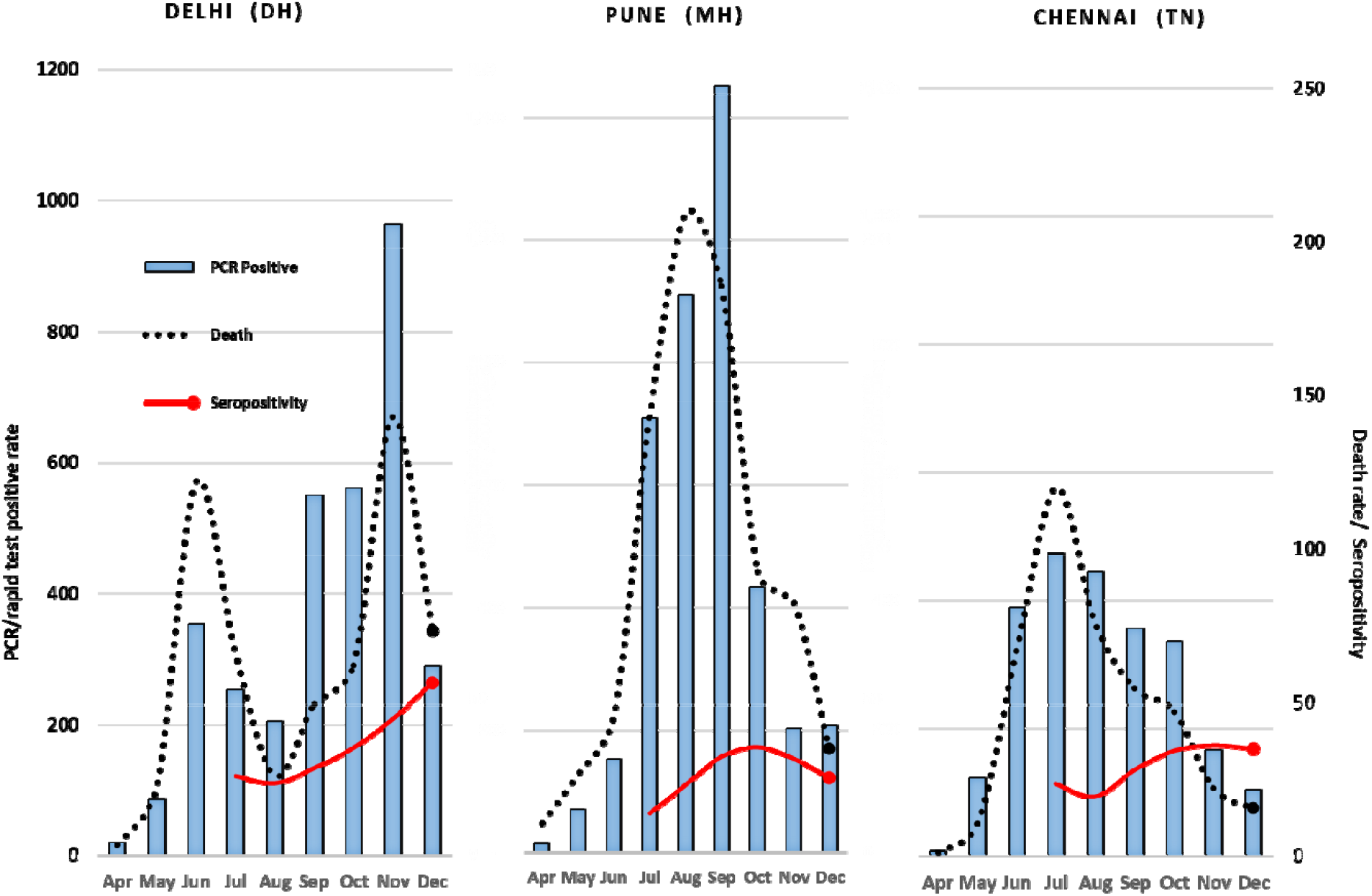
SARS-CoV-2 seropositivity (%), cases per 1,000 population and deaths per 100,000 population by month, selected cities. Left y-axis is the PCR/Rapid test positive rate per 100,000 population. Righty-axis applies to both % self-referred population IgG seropositivity and COVID death rate per 10,000 population.

An increase in measles vaccination among children in 2015-16 at the district level was associated with a marginally significant 6% reduction in seropositivity nationally (95%CI 0-12%), and there was a non-significant 2% reduced risk with increased polio vaccination (Figure 3). By contrast, PM2.5 per 10 unit increase, younger population structure or higher migrant history were each unassociated with increased seropositivity. The spatially-distributed risks of seropositivity at the pincode level varied, and at the state level, case and death rates peaked in most states in September-October (supplementary figure 5).

**Figure 3.**
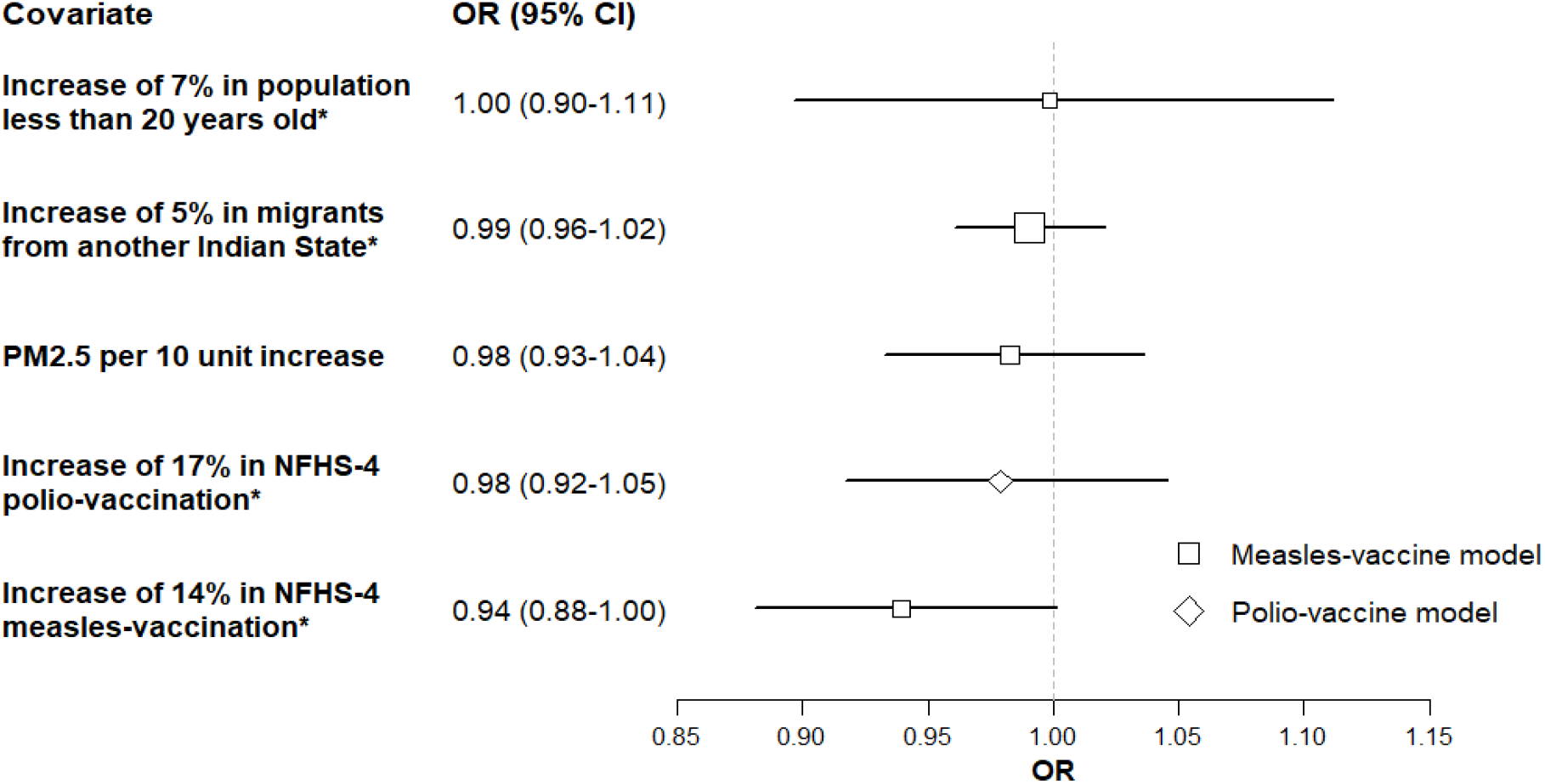
Risk factors for being a seropositive case. The number of seropositive serology testing included is 255,601. Covariates shown are sub-district level percentages (population less than aged 20, migrants from another state), mean PM2.5 within the Pincode area, and district-level NFHS-4 percentage of polio or measles vaccination. Symbol sizes are scaled to the inverse of the standard deviations of the estimates (i.e., larger standard deviation has smaller size). Abbreviations: OR odds ratio; CI credible interval. *The explanatory variables were standardized to either every unit increase in interquartile range for normally-distributed data, every unit increase in the difference between the 50th and (almost) 0th percentile for rightly skewed data, or every unit increase in the difference between the (almost) 100th and 50th percentile for left-skewed data. Thus, the units of increase for ORs are not the same across variables. This allows more direct comparison of effect size (Supplementary Appendix). The unstandardized means, medians, and ranges for each variable are as follows: Population less than 20 years old: mean 37%, median 35%, range 27-58%; Migrant: mean 11%, median 5%, range 0-59%; PM2.5: mean 70, median 53, range 27-134; NFHS-4 polio vaccination: mean 75%, median 76%, range 22-100%; NFHS-4 measles vaccination: mean 85%, median 86%, range 27-100%.

Because the tested individuals are not a representative sample of the urban Indian population, we also calculated mortality-based infection rates with an assumed infection fatality rate ranging from 0.29% to 0.21% based on independent sero studies in urban India, and adjusted COVID death totals in each city (Table 1, Figure 4, supplementary table 2). This suggests that overall in 12 cities about 43%-65% of adults above age 20 experienced SARS-CoV-2 infection in 2020, with a wide range of infection: Pune had the highest at 69% to an improbable 97%, whereas Jaipur had the lowest at 11%-15%. For these 12 cities, this suggests about 26 to 36 million adults above age 20 years had been infected. This corresponds to about 9 to 12 infected adults for every confirmed case, with variability in this ratio across cities (most confirmed cases were >20 years^3^). The mortality-derived prevalence varied considerably in the hypothetical scenarios of zero or 50% undercounts of COVID deaths (supplementary table 2). A subset of five cities had SARS-COV-2 antibody serosurveys on more randomly-selected populations, but there were no proper nationally-representative serosurveys. (Figure 4). These mostly showed a similarly high level of seropositivity (albeit with a wide range from 4-56%) and broadly similar patterns to mortality-derived prevalence (supplementary figure 6).

**Figure 4.**
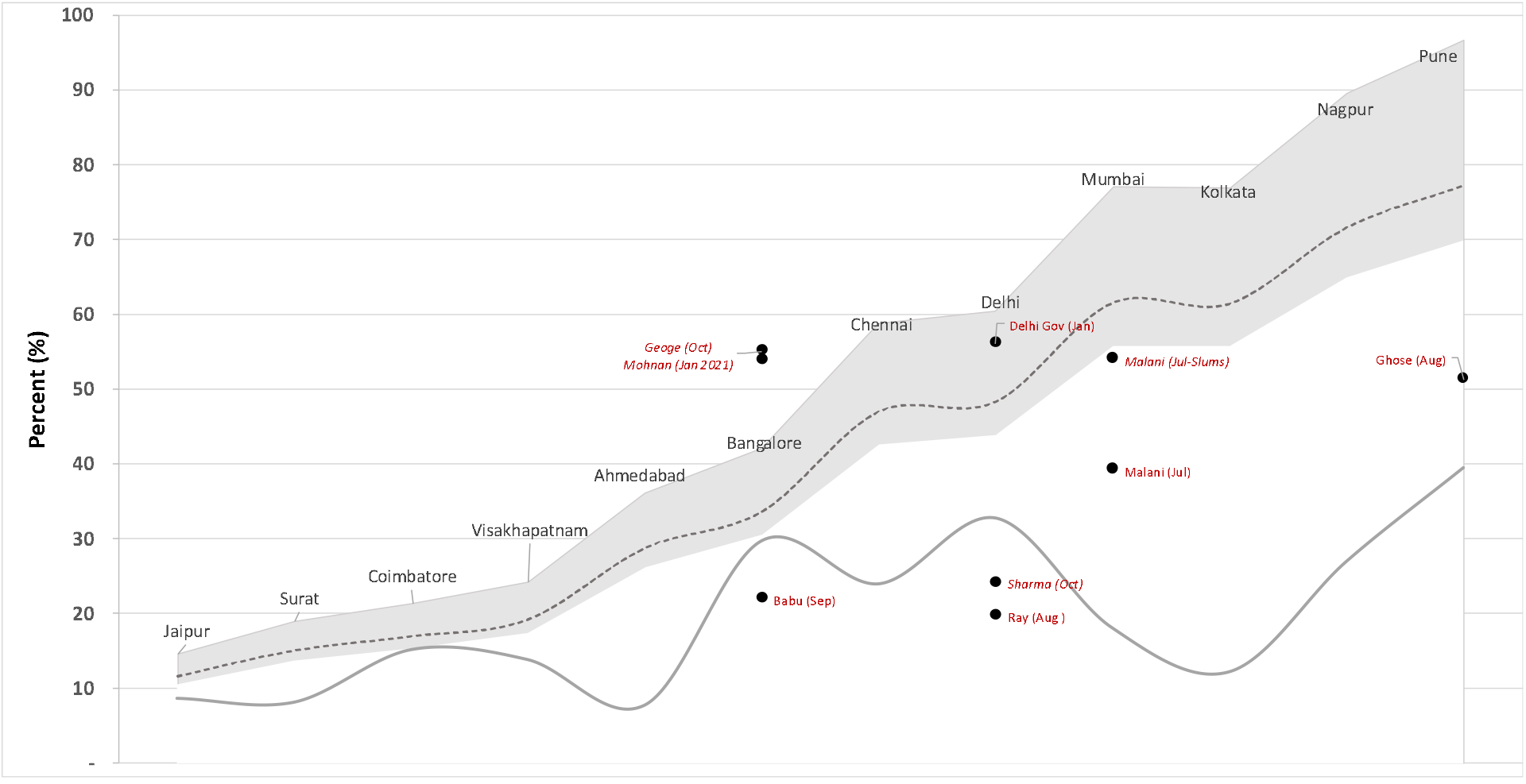
Mortality-derived prevalence and PCR confirmed test rate by city for Apr-Dec period. The grey bands represent the mortality-based prevalence using upper (0.21) and lower (0.29) infection fatality rates (IFR). The dotted curve in gray band represents middle prevalence correspondent to IFR=0.26 (see Table 1). The solid line represents PCR/antigen testing per 1000 population. The solid circles the seroprevalence in independent studies with more than 500 people tested with the labels showing the lead author and month of study.

## Discussion

We found a remarkably high seropositivity rate to SARS-CoV-2, approaching half of all adults in self-referred populations in selected major cities in India. The results were broadly consistent with the high prevalence estimated from COVID mortality data, as well as with a limited number of serological surveys done in more randomly-selected populations.

India’s urban COVID pandemic has been characterized as a paradox of widespread infection but low mortality.^8,24,25^ Urban India still has a young age structure and young adults have low IFRs in nearly all countries.^24,25^ The age-specific and temporal patterns suggesting substantial intergenerational transmission are consistent with Indian household composition. Similar seropositivity patterns by age are seen elsewhere where multiple generations live in one household, such as in Brazil and Iran.^26,27^ The most rapid increases in seropositivity through December at younger ages (<20) are consistent with younger cohorts in India being important transmitters of infection to older household members. The lowest OR of infection was at ages 20-24 years, when young adults are most likely to be living away from multigenerational households.^28^

Asymptomatic infection is more commonly reported in Indian serosurveys, exceeding 90% in some,^6,29^ in contrast to high-income countries, where about one-third of infections are asymptomatic.^30,31^ Asymptomatic infection may be less transmissible than from symptomatic index cases, and lead those infected also having milder disease.^32^ In addition, there has been speculation that non-specific effects of measles, polio, and BCG vaccination (each use live, attenuated viruses or bacteria) in the population may reduce the probability of SARS-CoV-2 infection or its severity.^33,34^ While our finding of reduced seropositivity with earlier childhood measles vaccine is based only on ecologic associations, it suggests that background immunity factors might help explain widespread infection and low mortality rates in India.

The alternative explanation, that urban India has a larger number of hidden deaths is certainly true in some settings, such as in the cities of Bihar and Uttar Pradesh, which have low levels of routine civil registration of deaths. However, we restricted our analyses only to the dozen cities with good registration in recent years,^16^ and hospitalization data from these cities^3^ show broadly similar levels and trends to COVID deaths. Under-reporting of COVID deaths of about 30-40% is now documented worldwide,^2,19^ and we incorporated such levels of under-reporting in our analyses. However, accounting for under-reporting resulted in even higher and sometimes implausibly high mortality-derived seroprevalence (supplementary table 2).

Widespread SARS-CoV-2 infection in urban India has several implications. First, subsequent viral waves remain possible. Indeed, Delhi and some other cities have had at least two viral waves and as of early March, modest increases in case counts were reported in various cities.^3^ Repeat waves could be expected with indoor crowding in many parts of India during April to June with extreme heat driving more people to cooler indoor settings. Second, the delivery of COVID vaccines practicable for widespread use in India would need to consider if immunity is already established, and the age-specific differences in durability of vaccine versus naturally-acquired immunity.^35^ Third, the reasons for the widespread variation in infection, including the role of cellular immunity, needs exploration.^36^

Notwithstanding the self-selected nature of the serosurvey population, our study has several advantages. The self-referred populations are likely biased to those seeking diagnosis and higher SES.^13^ Thus, the actual prevalence may be even greater, as serological studies in focal areas find that higher SES have lower prevalence than the lower SES.^6-9^ However, these biases were unlikely to have changed over the six-month study period. We capture the diverse regions of India, using standardized antibody assays with similar performance. Local mortality data enable a city-specific prevalence estimate, including if low prevalence was a consequence of under-reporting of deaths. Nonetheless, the study has some limitations. First, we could not provide reliable estimates of COVID infection or mortality in rural areas, where most of India’s 10 million annual deaths occur.^4^ In rural areas, SARS-CoV-2 infection and deaths might already be substantial. The Registrar General of India needs to re-start its Million Death Study,^4,37^ a mortality surveillance that provides cause of death information in over 8000 randomly-selected villages and urban blocks, as only maternal death data have been published after 2013. Repeat properly-representative large serosurveys that avoid some of the sampling limitations of current surveys^29^ are needed to monitor changes in natural and vaccine-induced immunity. Second, we had to rely on published estimates of IFR from three cities in Western India. IFRs may well vary by city and on variables such as background rates of obesity and other risk factors which are not well understood.^5^ The mortality-derived seroprevalence, while crude, is transparent and can be updated with improved IFR estimates.

## Conclusions

India’s COVID experience until early 2021 has been substantially different than that in high-income countries, with widespread intragenerational transmission. In every country, direct sero-epidemiological studies and improved COVID mortality reporting^38^ can document the path of the pandemic and assess the impact of vaccination and other control efforts.

## SUMMARY BOX

### What is already known on this topic

- Overall COVID mortality rates in urban India (where SARS-CoV-2 infection is concentrated) are substantially lower than those observed in high-income countries.
- Previous reporting on the seroprevalence of SARS-CoV-2 antibodies has been in focal urban areas, and show infection to be common.

### What this study adds

- For the whole of India, 31% of the self-referred people tested in over 2,200 collection points tested seropositive for SARS-CoV-2 antibodies using centralized assays and by December, nearly half of the tested population were seropositive.
- We observed higher seropositivity in females (35%) than in males (30%) in nearly every age group, and an “M-shaped” age pattern of seropositivity broadly consistent with widespread intergenerational transmission.
- Seropositivity was reduced in areas with higher childhood measles vaccination, perhaps reflecting non-specific effects on SARS-CoV-2 immunity.
- Based on local mortality data in the 12 cities, we estimate that 43%-65% of adults above age 20 were infected, corresponding 26 to 36 million adults in these cities, or an average of 9 to 12 infected per confirmed case.

## Data Availability

The data analysis code is available from the corresponding author.

## Acknowledgements

We thank Thyrocare Laboratories for providing the anonymized data to enable this analyses.

## Contributors

Conceived the idea and developed the study design: AV, CN, PJ. Data analysis: WS, SHF, PB, Literature review: WS, HG. PJ and AV wrote the initial draft, and all authors were involved in commenting on subsequent revisions. The corresponding author attests that all listed authors meet authorship criteria and that no others meeting the criteria have been omitted.

## Ethical approval

The study used anonymized data with no personal identifiers. Ethical approval was provided under an overall IRB for use of anonymized mortality and serosurveillance data by Unity Health REB, Toronto.

## Data sharing

The data analysis code is available from the corresponding author.

The corresponding author (PJ) affirms that the manuscript is an honest, accurate, and transparent account of the study being reported; that no important aspects of the study have been omitted; and that any discrepancies from the study as planned (and, if relevant, registered) have been explained.

Dissemination to participants and related patient and public communities: Direct dissemination to study participants is not possible. Preprint versions of this analysis are available on MedRxiv.

## References

1. Coronavirus.app. The Coronavirus App [Internet]. Taipei, Taiwan: Coronavirus.app. 2020 [cited 2021 Jan 3];Available from https://coronavirus.app/map

2. Brown P, Rai K, La Vecchia C, et al. Mortality from COVID-19 in 12 countries and 6 states of the United States. medRxiv 2020. doi: https://doi.org/10.1101/2020.04.17.20069161. Available from https://covid.cghr.dev/.

3. COVID-19 India Org Data Operations Group. A volunteer-driven, crowd-sourced database for COVID-19 stats & patient tracing in India [Internet]. 2020 [cited 2021 Jan 05];Available from https://api.covid19india.org/

4. Menon GR, Singh L, Sharma P et al. National Burden Estimates of healthy life lost in India, 2017: an analysis using direct mortality data and indirect disability data. Lancet Glob Health. 2019 Dec;7(12):e1675–e1684. doi: 10.1016/S2214-109X(19)30451-6.

5. Walker PG, Whittaker C, Watson OJ, et al. The impact of COVID-19 and strategies for mitigation and suppression in low-and middle-income countries. Science. 2020 Jun 12.

6. George CE, Inbaraj LR, Chandrasingh S, de Witte LP. High seroprevalence of COVID-19 infection in a large slum in South India; what does it tell us about managing a pandemic and beyond? Epidemiology and Infection. 2021;149, e39. 1–6. doi: https://doi.org/10.1017/S0950268821000273

7. Malani A, Shah D, Kang G, et al. Seroprevalence of SARS-CoV-2 in slums versus non-slums in Mumbai, India [published online ahead of print, 2020 Nov 13]. Lancet Glob Health. 2020;S2214-109X(20)30467-8. doi:10.1016/S2214-109X(20)30467-8

8. Ghose A, Bhattacharya S, Karthikeyan AS, et al. Community prevalence of antibodies to SARS-CoV-2 and correlates of protective immunity in five localities in an Indian metropolitan city. medRxiv 2020 Dec. doi: https://doi.org/10.1101/2020.11.17.20228155

9. Mohanan M, Malani A, Krishnan K, et al. Prevalence of SARS-CoV-2 in Karnataka, India. JAMA. 2021;325(10):1001–1003. doi: 10.1001/jama.2021.0332

10. International Institute for Population Sciences (IIPS) and ICF. National Family Health Survey (NFHS-4), 2015-16: India. 2017. Mumbai: IIPS.

11. Gupta A, Bherwani H, Gautam S, et al. Air pollution aggravating COVID-19 lethality? Exploration in Asian cities using statistical models [published online ahead of print, 2020 Jul 15]. Environ Dev Sustain. 2020;1-10. doi:10.1007/s10668-020-00878-9

12. Kallathiyan K, Velumani A, Iyer S, Sivapandi K, Velumani A. COVID-19 seroprevalence study of an Indian Diagnostic Laboratory-Report on gender and age analysis. Asian Journal of Health Sciences. 2020 Nov 10;6(2):16.

13. Sahasranaman A, Kumar N. Income distribution and inequality in India: 2014-19. arXiv preprint 2010.03602. 2020 Oct 7.

14. Abbott Laboratories. Abbott ARCHITECT SARS-CoV-2 IgG Reagent Instructions for Use [Internet]. December 2020 [cited 2020 Dec 01]; Available from: https://www.fda.gov/media/137383/download

15. Roche. Elecsys® Anti-SARS-CoV-2 [Internet]. 2020 [cited 2020 Dec 01];Available from: https://diagnostics.roche.com/global/en/products/params/elecsys-anti-sars-cov-2.html.

16. Registrar General of India. 2018. Vital Statistics of India Based on the Civil Registration System 2018. New Delhi: Office of the Registrar General of India, Ministry of Home Affairs, Government of India.

17. World Health Organization. Emergency use ICD codes for COVID-19 disease outbreak [Internet]. 2020 [cited 2021 Jan 4]; Available from https://www.who.int/standards/classifications/classification-of-diseases/emergency-use-icd-codes-for-covid-19-disease-outbreak

18. Government of India. Covid-19 Update [Internet]. 2020 [cited 2020 Dec 01]; Available from: https://www.mygov.in/covid-19/

19. Overberg P, Kamp J, Michaels D, Huth L. The Covid-19 Death Toll Is Even Worse Than It Looks [Internet]. Wall Street Journal. January 15, 2021 [cited 2021 Jan 17]; Available from https://www.wsj.com/articles/the-covid-19-death-toll-is-even-worse-than-it-looks-11610636840

20. GfK GeoMarketing GmbH. GfK new digital maps for Asia [Internet]. 2018 [cited 2021 Jan 16]; Available from https://www.gfk.com/fileadmin/user_upload/dyna_content/Global/documents/Press_Releases/2018/20180321_PR_GfK_Asia-map-edition_efin.pdf

21. Hammer MS, van Donkelaar A, Li C, et al. Global Estimates and Long-Term Trends of Fine Particulate Matter Concentrations (1998-2018). Environmental Science & Technology. 2020 Jun 3. doi: https://pubs.acs.org/doi/10.1021/acs.est.0c01764

22. WorldPop and Center for International Earth Science Information Network (CIESIN), Columbia University (2018). Global High Resolution Population Denominators Project. doi: https://dx.doi.org/10.5258/SOTON/WP00649

23. Brown PE. 2015. Model-based geostatistics the easy way. Journal of Statistical Software 63: 1–24.

24. Levin AT, Hanage WP, Owusu-Boaitey N, Cochran KB, Walsh SP, Meyerowitz-Katz G. Assessing the age specificity of infection fatality rates for COVID-19: systematic review, meta-analysis, and public policy implications. European journal of epidemiology. 2020 Dec 8:1–6.

25. O’Driscoll M, Ribeiro Dos Santos G, Wang L, et al. Age-specific mortality and immunity patterns of SARS-CoV-2. Nature 2020. https://doi.org/10.1038/s41586-020-2918-0

26. Hallal PC, Hartwig FP, Horta BL, et al. SARS-CoV-2 antibody prevalence in Brazil: results from two successive nationwide serological household surveys. The Lancet Global Health. 2020 Nov 1;8(11):e1390–8.

27. Poustchi H, Darvishian M, Mohammadi Z, et al. SARS-CoV-2 antibody seroprevalence in the general population and high-risk occupational groups across 18 cities in Iran: a population-based cross-sectional study. Lancet Infect Dis. 2020;S1473-3099(20)30858-6. doi:10.1016/S1473-3099(20)30858-6

28. Dommaraju P. One-person households in India. Demographic Research. 2015 Jan 1;32:1239-66.

29. Murhekar MV, Bhatnagar T, Selvaraju S, et al. SARS-CoV-2 antibody seroprevalence in India, August– September, 2020: findings from the second nationwide household serosurvey. The Lancet Global Health. 2021 Mar 1;9(3):e257–66.

30. Buuitrago-Garcia D, Egli-Gany D, Counotte MJ, et al. Occurrence and transmission potential of asymptomatic and presymptomatic SARS-CoV-2 infections: A living systematic review and meta-analysis. PLoS Med. 2020;17(9):21003346

31. Byambasuren O, Cardona M, Bell K, Clark J, McLaws ML, Glasziou P. Estimating the extent of asymptomatic covid-19 and its potential for community transmission: Systematic review and meta-analysis, Off. J. Assoc. Med. Microbiol. Infect. Dis. Canada COVID-19, e20200030 (2020)

32. Sayampanathan AA, Heng CS, Pin PH, et al. Infectivity of asymptomatic versus symptomatic COVID-19. The Lancet. 2021 Jan 9;397(10269):93–4.

33. Aaby P, Benn CS, Flanagan KL, et al. The non-specific and sex-differential effects of vaccines. Nature Reviews Immunology. 2020 Aug;20(8):464–70.

34. Chumakov K, Benn CS, Aaby P, Kottilil S, Gallo R. Can existing live vaccines prevent COVID-19?. Science. 2020 Jun 12;368(6496):1187–8.

35. Lumley SF, O’Donnell D, Stoesser NE, et al. Antibody status and incidence of SARS-CoV-2 infection in health care workers. New England Journal of Medicine. 2021 Feb 11;384(6):533–40.

36. Sewell HF, Agius RM, Stewart M, Kendrick D. Cellular immune responses to covid-19. BMJ 2020; 370:m3018.

37. Menon GR, Singh L, Sharma P, et al. National Burden Estimates of healthy life lost in India, 2017: an analysis using direct mortality data and indirect disability data. Lancet Glob Health. 2019;7(12):e1675–e1684. doi:10.1016/S2214-109X(19)30451-6

38. Lipsitch M, Swerdlow DL, Finelli L. Defining the Epidemiology of Covid-19 – Studies Needed. New England Journal of Medicine. 2020;382(13):1194—1196.

