## Supplementary appendix for "SARS-CoV-2 Seroprevalence in 12 Cities of India from July-December 2020"

**Supplementary Appendix to: *SARS-CoV-2 Seroprevalence, Cases, and Mortality in 12 Cities of India from July-December 2020* A Velumani et al**

**Contents**

Supplementary Statistical Methods	2

Supplementary tables 1-3	5

Supplementary figures 1-7	8

References	14

### Supplementary Statistical Methods

### Introduction

The statistical models were fitted using Bayesian inference, and posterior distributions were computed using the INLA methodology from inla (Rue et al., 2009), as explained in the following sections.

#### **Spatially-smoothed odds ratio of SARS-CoV-2 seropositivity**

A logistic regression model with a spatial random effect was used for the spatial analyses of seropositivity, using a model form of Generalized Linear Geostatistical Model described by Diggle & Ribeiro (2006).

Writing $Y_{ij}$ as the number of seropositive cases in Pincode area $i$ (located at the spatial coordinates $s_{i}$) for age group $j$, the model is as follows:

$$Y_{ij}\sim\text{Binom}\left( n_{ij}{,p}_{ij} \right)$$

$$logit\left( p_{ij} \right)=X\left( s_{i} \right)\alpha+f\left( A_{j} \right)+U\left( s_{i} \right)+Z_{i}$$

$$f\left( A \right)\sim RW2\left( \gamma\right)$$

$$Z_{i}\sim N\left( 0,\tau^{2} \right)$$

$$U\left( s \right)\sim N\left( 0,\sigma^{2} \right)$$

$$\text{cor}\left[ U\left( s+h \right),U\left( s \right) \right]=\sigma^{2}\rho\left( \left| \left| h \right| \right|/\phi\right)$$

$n_{ij}$ is the number patients undergone the serology testings. The log odds ratio (i.e., logit-transformed of $p_{ij}$) depends on the spatially referenced covariates $X\left( s_{i} \right)$, the spatial random effect $U\left( s_{i} \right)$, and pincode unit-level effect $Z_{i}$.

- The spatial explanatory variable $X\left( s_{i} \right)$ includes an intercept; a sex indicator; percentage of urban settlements, population size, and population density in the pincode area for 2020; particulate matter less than 2.5 microns per cubic meter (μg/m^3^) (PM_2.5_) in the pincode area, using mean value of 2015-2017 (Hammer et al., 2020); sub-district-level proportion of female illiteracy, migrants from another Indian State, population less than 20 years old, solid-fuel use, and dominant language groups; and percentage of children age 12-23 months who have received measles vaccines (or polio vaccines) at the district-level from NFHS-4 (IIPS and ICF, 2017), linked to the pincode areas. We derived these covariates mostly from 2011 Indian census or WorldPop gridded data (WorldPop and CIESIN, 2018).
- Except for dominant language group and PM_2.5_, the explanatory variables were standardized to either every unit increase in interquartile range for normally-distributed data, every unit increase in the difference between the 50^th^ and 0^th^ percentile for rightly skewed data, or every unit increase in the difference between the 100^th^ and 50^th^ percentile for leftly skewed data. This method of standardization allows more directly comparison of the effect size (of odds ratio) between the covariates, as each covariate has different data range.
- $f\left( A_{j} \right)$ is the log-odds ratio for age group $A_{j}$, modelled as a second order random walk with standard deviation $\gamma$.
- The spatial random effect $U\left( s_{i} \right)$ is a Gaussian random field with a Matern spatial correlation function $\rho$ specifying how correlation decreases with distance $h$, depending on the value of a range parameter $\phi$. The range parameter determines how rough or smooth the relative risk surface is.
- The unit-level effect $Z_{i}$ is spatially independent, and unlike the spatial effect $U\left( s_{i} \right)$, two sampling units in close proximity will have unrelated values of $Z_{i}$. The $Z_{i}$ can be thought of as accounting for short-scale spatial variation or pincode-unit-level risk factors not included in the model as covariates.

The smoothed odds ratio mapped in web figure 3C is the posterior median of the spatial odds ratio $exp\left[ X\left( s \right)\alpha+U\left( s \right) \right]$, where $X$ exclude the percentage of urban settlements, population size, and population density due to their gridded data are too huge for calculating the predictions. Thus, the smoothed odds ratios represent zero value for these variables, while accounting for the spatially varying values for the other explanatory variables (e.g. PM_2.5_, female illiteracy).

To obtain the non-linear effect of age by sex (Figure 1B), we ran separate models for men and women to obtain the estimates.

We fit separate spatial and temporal models as a combined spatio-temporal model would be unfeasible due to computational requirements. See section below on specifications for the temporal models.

##### ***Implementation***

As there are 1,415 geocoded pincode areas and spatial locations $s_{i}$, model fitting is computationally intensive and an approximation to the spatial covariance matrix is necessary. The Markov random field approximation from Lindgren et al. (2011) is used here, and implemented in the geostatsp package (Brown, 2015) for the R statistical programming language (R Core Team, 2018), which in turns calls the inla software (Rue et al., 2009). A full description of the methodology can be found in Brown (2016). Although there are other methods available for fitting models of this type, the task is complex and computationally demanding and there is currently no rival to the Bayesian methodology in the inla software for fitting spatial models with non-Normal responses.

Bayesian inference requires specifying prior distributions, and spatial models are particularly susceptible to producing spurious results from ill-chosen priors for the spatial parameters $\phi$ and $\sigma$. Here the penalized complexity prior distributions from Simpson et al. (2017) are used, priors which discourage a spatial effect (wanting $U\left( s \right)$ flat and close to zero) unless the data indicate a clear preference for a spatial model. Following Simpson et al. (2017), the standard deviations , $\sigma$ and $\tau$ have exponential priors, as does the scale parameter $1/\phi$. The prior median for $\phi$, the distance beyond which the correlation between two locations is under 10%, is $500km$ or roughly one sixth of the distance across India. The prior medians of $\sigma$ and $\tau$ are both $\text{log}\left( 2 \right)$, a value at which a one standard deviation increase in $U\left( s_{i} \right)$ or $Z_{i}$ doubles mortality risk.

##### ***Parameter estimates***

Here we provide the parameter estimates for covariates, spatial parameters, and hyperparameters for random effects included in the Bayesian spatial logistic model. We report the median (50^th^ percentile), 2.5^th^ and 97.5^th^ percentile of the posterior distributions.

|  | **Odds ratio of seropositivity** | | |
| --- | --- | --- | --- |
|  | **Median** | **2.5^th^ pct** | **97.5^th^ pct** |
| (Intercept) | 0.395 | 0.273 | 0.550 |
| Sex: Female (ref Male) | 1.112 | 1.095 | 1.130 |
| Percentage of urban areas | 1.001 | 0.984 | 1.017 |
| Standardized female illiteracy | 1.009 | 0.911 | 1.118 |
| PM 2.5 per 10 unit increase | 0.983 | 0.933 | 1.036 |
| Standardized solid-fuel use | 0.975 | 0.885 | 1.074 |
| Language: Dravidian (reference) |  |  |  |
| Language: Hindi | 1.151 | 0.803 | 1.663 |
| Language: Austroasiatic | 0.794 | 0.155 | 4.056 |
| Language: Magadhan | 1.200 | 0.837 | 1.738 |
| Language: Marathi-Konkani | 1.064 | 0.797 | 1.433 |
| Language: Old Gujarati | 1.060 | 0.728 | 1.561 |
| Language: Pahari-Dardic | 1.276 | 0.692 | 2.379 |
| Language: Punjabi | 1.202 | 0.724 | 1.987 |
| Population size | 0.981 | 0.972 | 0.990 |
| Population density | 1.019 | 1.009 | 1.029 |
| Pct less than 20 years old | 0.999 | 0.897 | 1.112 |
| Pct migrants from another Indian state | 0.990 | 0.961 | 1.021 |
| Pct of children age 12-23 months who have received measles vaccines | 0.939 | 0.881 | 1.001 |
| SD for age effect (γ) | 0.008 | 0.006 | 0.012 |
| SD for Pincode unit effect (τ) | 0.378 | 0.352 | 0.406 |
| SD of spatial random effect (σ) | 0.372 | 0.303 | 0.477 |
| range/1000 ($\phi$/1000) | 16.659 | 11.104 | 27.594 |

SD: standard déviation, pct: percentile

#### **Temporal models for SARS-CoV-2 seropositivity and Covid-19 cases and deaths**

We explore the temporal effects using a non-spatial logistic regression to model the odds ratio of seropositivity prevalence in reference to July 1st, using second order random walks to model the non-linear temporal effects. The logistic regression model also included an age effect (categorized to five-year age groups and ages 80+) and day-of-week effect.

We converted the model estimates from the logistic regression to absolute daily seropositivity prevalence by this formula: $\sum_{j} \mathrm{invlogit}\left( \beta_{0}+ {RW2}_{date}+a_{j} \right)\times{total no. of serology tests}_{j}$, where $\beta_{0}$ is the model intercept, ${RW2}_{date}$ represents the log odds ratios for each date as estimated from second order random walks, $a$ is the log odds ratio for each age category, and $j$ represents age group. The daily seropositivity prevalence were further converted to daily number of new seropositive cases by taking the difference of prevalence between consecutive days.

We used similar methods to explore the relative risks of Covid cases and deaths in reference to May 1^st^. We used non-spatial Poisson regressions instead of logistic regression. The Poisson regression models also included a day-of-week effect. The model estimates were converted to absolute daily cases/deaths using this formula: $\exp\left( \beta_{0}+ {RW2}_{date} \right)\times UN estimated 2020 Indian population$, where $\beta_{0}$ is the model intercept, ${RW2}_{date}$ represents the log relative risks for each date as estimated from second order random walks.

**Supplementary Table 1 – Demographic characteristics and anti-SARS-CoV-2 IgG seroprevalence at periodic testing in most populated cities from July to December 2020**

| Characteristic |  | | Total Jun-Dec  (% total) | | Jun | | Jul | | Aug | | Sep | | Oct | | Nov | | Dec |
| --- | --- | --- | --- | --- | --- | --- | --- | --- | --- | --- | --- | --- | --- | --- | --- | --- | --- |
| Total tested | Total | | 448,518 (100%) | | 5,298 | | 68,474 | | 117,438 | | 116,395 | | 73,831 | | 40,482 | | 26,600 |
|  | % Positive | | 31.4% | | 5.4% | | 16.0% | | 26.0% | | 31.9% | | 41.3% | | 45.8% | | 48.0% |
| Gender |  | |  | |  | |  | |  | |  | |  | |  | |  |
| Male | Total | | 308,958 (68.9%) | | 4,362 | | 49,716 | | 83,860 | | 81,531 | | 48,036 | | 24,825 | | 16,628 |
|  | % Positive | | 29.9% | | 4.8% | | 15.0% | | 25.0% | | 30.5% | | 40.8% | | 45.7% | | 47.1% |
| Female | Total | | 139,560 (31.1%) | | 936 | | 18,758 | | 33,578 | | 34,864 | | 25,795 | | 15,657 | | 9,972 |
|  | % Positive | | 34.6% | | 8.2% | | 18.7% | | 28.3% | | 35.0% | | 42.3% | | 45.9% | | 49.5% |
| Age in years | Median years | | 37 | | 33 | | 36 | | 36 | | 37 | | 38 | | 40 | | 40 |
| <20 | Total | | 23,339 (5.2%) | | 144 | | 3,785 | | 5,691 | | 6,028 | | 3,960 | | 2,281 | | 1,450 |
|  | % Positive | | 32.0% | | 4.9% | | 15.7% | | 27.3% | | 32.8% | | 41.7% | | 44.8% | | 46.0% |
| 20-44 | Total | | 274,288 (61.2%) | | 4,072 | | 43,780 | | 75,714 | | 71,827 | | 42,485 | | 21,944 | | 14,466 |
|  | % Positive | | 27.8% | | 4.8% | | 14.1% | | 23.5% | | 28.7% | | 38.0% | | 41.6% | | 43.7% |
| 45-69 | Total | | 135,627 (30.2%) | | 1,040 | | 19,078 | | 32,636 | | 34,795 | | 24,392 | | 14,320 | | 9,366 |
|  | % Positive | | 37.5% | | 7.8% | | 19.8% | | 30.9% | | 37.6% | | 46.6% | | 51.6% | | 53.9% |
| 70 or above | Total | | 15,264 (3.4%) | | 42 | | 1,831 | | 3,397 | | 3,745 | | 2,994 | | 1,937 | | 1,318 |
|  | % Positive | | 39.1% | | 14.3% | | 22.8% | | 32.7% | | 37.1% | | 45.4% | | 50.1% | | 54.4% |
| Single or dual Assay | |  | |  | |  | |  | |  | |  | |  | |  | |
| Abbott alone | Total | | 218,111 (48.6%) | | - | | 13,034 | | 66,694 | | 63,796 | | 39,424 | | 23,083 | | 12,080 |
|  | % Positive | | 30.5% | | - | | 17.1% | | 24.0% | | 29.6% | | 38.4% | | 40.5% | | 41.7% |
| Roche alone | Total | | 103,221 (23.0%) | | 5,298 | | 43,139 | | 13,642 | | 18,739 | | 11,926 | | 5,121 | | 5,356 |
|  | % Positive | | 24.5% | | 5.4% | | 14.1% | | 24.0% | | 27.8% | | 43.0% | | 50.5% | | 50.7% |
| Both agree | Total | | 118,499 (26.4%) | | - | | 11,745 | | 35,446 | | 31,912 | | 20,961 | | 10,508 | | 7,927 |
|  | % Positive | | 27.1% | | . | | 17.9% | | 27.1% | | 34.5% | | 41.5% | | 46.0% | | 47.6% |
| Discordant | Total | | 8,687 (1.9%) | | - | | 556 | | 1,656 | | 1,948 | | 1,520 | | 1,770 | | 1,237 |
|  | % positive Abbott | | 31.5% | |  | | 60.4% | | 51.8% | | 41.6% | | 30.3% | | 10.9% | | 6.5% |
|  | % positive Roche | | 68.5% | |  | | 39.6% | | 48.2% | | 58.4% | | 69.7% | | 89.1% | | 93.5% |
| 3 mega cities | Total | | 99,569 (22.2%) | | 402 | | 12,766 | | 28,095 | | 21,459 | | 17,600 | | 11,631 | | 7,616 |
|  | % Positive | | 33.3% | | 24.9% | | 24.8% | | 26.1% | | 31.1% | | 38.6% | | 46.9% | | 48.1% |
| 9 other cities | Total | | 85,702 (19.1%) | | 1,529 | | 12,301 | | 26,300 | | 20,656 | | 13,893 | | 6,469 | | 4,554 |
|  | % Positive | | 27.3% | | 1.7% | | 15.2% | | 24.1% | | 28.7% | | 35.6% | | 38.2% | | 40.0% |
|  | No. tested/ 100,000 person | | 201 | | 4 | | 29 | | 62 | | 48 | | 33 | | 15 | | 11 |
| All other areas | Total | | 263,247 (58.7%) | | 3,367 | | 43,407 | | 63,043 | | 74,280 | | 42,338 | | 22,382 | | 14,430 |
|  | % Positive | | 31.9% | | 4.8% | | 13.7% | | 26.7% | | 32.9% | | 44.3% | | 47.4% | | 50.5% |

**Supplementary Table 2 – SARS-Cov-2 mortality based seroprevalence sensitivity estimates in 12 cities in India**

|  |  |  | | | | Mortality-derived seroprevalence (%) | | | |  | |
| --- | --- | --- | --- | --- | --- | --- | --- | --- | --- | --- | --- |
| City (state, and population in millions) | | **Percent completeness of civil death registration 2018** * | **Overall deaths>**  **20 years in 2019 (pre-COVID) in 000** | **Death rate per 000 population in 2019 (pre-COVID)** | **Adjusted COVID proportion of deaths 2020 (%)** | **High IFR (0.29%)** | **Low IFR**  **(0.21%)** | **Sensitivity analyses** | | **Peak sero-positivity** | **Ratio of mortality-based prevalence to confirmed cases**  **High, low** † |
|  |  |  |  |  |  |  |  | **No undercount**  **High, low** | **50% undercount**  **High, low** |  |  |
| Mumbai (MH,18.4) | | 98.4 | 104.1 | 7.0 | 15.5 | 55.7 | 77.0 | 44.9 , 62.0 | 62.5 , 86.3 | 41.3 | 12.4 , 17.1 |
| Delhi (DH,16.3) | | 100.0 | 57.7 | 5.6 | 19.2 | 43.8 | 60.5 | 35.2 , 48.7 | 49.1 , 67.8 | 54.9 | 7.2 , 10.0 |
| Kolkata (WB,14.1) | | 91.8 | 41.4 | 8.2 | 15.8 | 55.6 | 76.8 | 33.8 , 46.6 | 48.2 , 66.6 | 54.6 | 10.2 , 14.0 |
| Chennai (TN,8.7) | | 100.0 | 45.1 | 8.6 | 12.9 | 42.6 | 58.8 | 24.0 , 33.2 | 34.7 , 47.9 | 36.2 | 7.5 , 10.4 |
| Bangalore (KN,8.5) | | 100.0 | 52.2 | 7.9 | 10.3 | 30.5 | 42.1 | 20.5 , 28.3 | 29.7 , 41.0 | 42.3 | 4.8 , 6.6 |
| Ahmedabad (GJ,6.4) | | 100.0 | 30.1 | 7.6 | 9.3 | 26.1 | 36.1 | 69.9 , 96.5 | 69.9 , 96.5 | 48.7 | 15.5 , 21.5 |
| Pune (MH,5.1) | | 98.4 | 27.2 | 7.0 | 22.2 | 69.9 | 96.5 | 10.7 , 14.8 | 15.7 , 21.7 | 34.5 | 7.3 , 10.1 |
| Surat (GJ,4.6) | | 100.0 | 23.6 | 7.6 | 5.1 | 13.7 | 19.0 | 8.3 , 11.4 | 12.2 , 16.8 | 47.9 | 8.7 , 12 |
| Jaipur (RJ,3.1) | | 99.9 | 15.4 | 7.4 | 4.1 | 10.6 | 14.7 | 52.5 , 72.5 | 72.4 , 99.0 | 63.7 | 3.9 , 5.4 |
| Nagpur (MH,2.5) | | 98.4 | 14.9 | 7.0 | 21.8 | 64.8 | 89.5 | 12.0 , 16.6 | 17.7 , 24.4 | 49.8 | 11 , 15.3 |
| Coimbatore (TN,2.2) | | 100.0 | 16.2 | 8.6 | 5.0 | 15.5 | 21.3 | 13.7 , 18.9 | 20.0 , 27.6 | 38.2 | 5.5 , 7.6 |
| Visakhapatnam (AP,1.7) | | 100.0 | 9.5 | 6.9 | 7.0 | 17.5 | 24.2 | 33.8 , 46.6 | 48.2 , 66.6 | 51.9 | 4.1 , 5.7 |
| *3 mega Cities (48.8)* | | **96.7** | **203.2** | **7.1** | 18.5 | 52.2 | 72.0 | 41.9 , 57.9 | 58.5 , 80.8 | **47.2** | 10.0 , 13.9 |
| *Total 12 cities (91.5)* | | **98.7** | **437.4** | **7.5** | 15.4 | 43.1 | 59.5 | 35.3 , 48.8 | 43.6 , 60.2 | **41.4** | 8.5 , 11.7 |

Notes:

SARS-Cov-2 PCR testing results and confirmed deaths from 26 April to December 31, 2020; * Office of Registrar General of India, Report of Vital Statistics of India based on the Civil registration System 2018 (Mahapatra, 2010); we applied an adjustment factor of 1.3 to confirmed COVID deaths in Table 1 to obtain the COVID proportional deaths and divided from sum of 2019 overall deaths and 2020 Covid deaths. Mortality-derived seroprevalence; high IFR 0.21 is drawn from Ghose et al., 2020 and lower IFR is from George et al., 2021. † For populations age 20 years or older, we do not adjust for the small percentage of cases below age 20.

**Supplementary Table 3 – Key features and results of selected urban population-based SARS-CoV-2 antibodies studies in India 2020**

| **First author (publication month in 2020)** | **Survey period** | **City/State** | **Setting** | **Serology test** | **No. tested** | **No. positive** | **Sero-prevalence** | **Asymptomatic** | **Infection fatality rate (IFR)** | **Matched seroprevalence** | |
| --- | --- | --- | --- | --- | --- | --- | --- | --- | --- | --- | --- |
|  |  |  |  |  |  |  |  |  |  | **In current study*** | **Mortality derived** |
| Kshatri (Aug) | Aug | Odisha | Urban households | Roche | 709 | 131 | 18% | 93% | n.a. | 17% |  |
| Satpati (Aug) | Jul -Aug | West Bengal | Urban households | Calbiotech | 458 | 19 | 4% | n.a. | n.a. | 19% |  |
| Malani (Sep) | Jun 29-19 | Mumbai, MH | Total | Abbott | 6904 | 2708 | 39% |  | 0.12% | 28% | 45-62% |
|  |  |  | Urban slums |  | 4202 | 2273 | 54% |  | 0.08% |  |  |
|  |  |  | Non-slums |  | 2702 | 435 | 16% |  | 0.26% |  |  |
| Khan (Sept) | Jul 1-15 | Jammu & Kashmir | Hospital visitors to 20 hospitals | Abbott | 2906 | 111 | 4% | n.a. | n.a. | 13% |  |
| Ray (Aug) | Jun-Aug | Delhi | Hospitalized patients | THSTI-RBD | 212 | 42 | 20% | n.a. | n.a. | 26% | 44-60% |
| Siddiqui (Sept) | Apr-Aug | Delhi | Hospital in Delhi | Roche | 780 | 152 | 19% | n.a. | n.a. | 22% | 44-60% |
|  |  |  | Staff |  | 448 | 74 | 17% |  |  |  |  |
|  |  |  | Visitors |  | 332 | 78 | 23% |  |  |  |  |
| ICMR Sero group (Nov) ± | Aug-Sep | 70 districts in 21 states | 70 districts | Abbott | 29,082 | 3135 | 7% | 98% | 0.09-0.11% | 33% | 31-43% |
|  |  |  | Urban (Slums) |  | 2,626 | 574 | 17% |  |  |  |  |
|  |  |  | Urban (Non slums) |  | 4,932 | 672 | 9% |  |  |  |  |
| Babu (Nov) | Sept 3-6 | Karnataka | General population | Kavach | 15,624 | 2565 | 27% | n.a. | 0.05% | 28% |  |
|  |  |  | Bangalore urban |  | 3,617 | n.a. | 22% |  | 0·10% |  | 30-42% |
| Ghose (Nov) | Jul -Aug | Pune, Maharashtra | High prevalence city wards | Eurimmun | 2,089 | 857 | 51% |  | 0.21% | 22% | 70-96% |
| George (Feb 2021) | Oct | Karnataka | Bangalore urban | Elecsys | 499 |  | 55% | 95% | 0.29% | 33% | 30-42% |
| Sharma (Dec) | Aug - Oct | Delhi | Urban | ERBALISA |  |  |  | n.a. | n.a. |  | 44-60% |
|  | Aug |  |  |  | 15,046 | 4267 | 28% |  | 0.08% | 23% |  |
|  | Sept |  |  |  | 17,409 | 4311 | 24% |  | 0.11% | . |  |
|  | Oct |  |  |  | 15,015 | 3829 | 24% |  | 0.13% | 34% |  |
| Gov. Delhi (Feb 2021) | Jan 2021 | Delhi | Urban | CLIA | 28,800 | n.a. | 56% | n.a. | n.a. | 55% | 44-60% |
| Mohanan (Jan 2021) | Jun-Aug 2020 | Karnataka | Urban | BioMérieux | 1,386 | n.a. | 53.8% | n.a. | n.a. | 20% |  |
| **Summary of studies** | **Apr 2020-Jan 2021** | **5 cities, 70 districts** | **Predominantly urban** | **-** | **176,502** | **28,123** | 3.6-56.1% | **93-98%** | **0.05-0.29%** | **13-55%** | **31-96%** |

Notes: * Matched for study month, location and study setting to the self-reported seropositivity data. Mortality derived seroprevalence uses the IFR of 0.21%-29% and the combination of states and time periods. ± We exclude first ICMR serosurvey done in May-June as that reported quite low seroprevalence and is mostly outside the study period here.

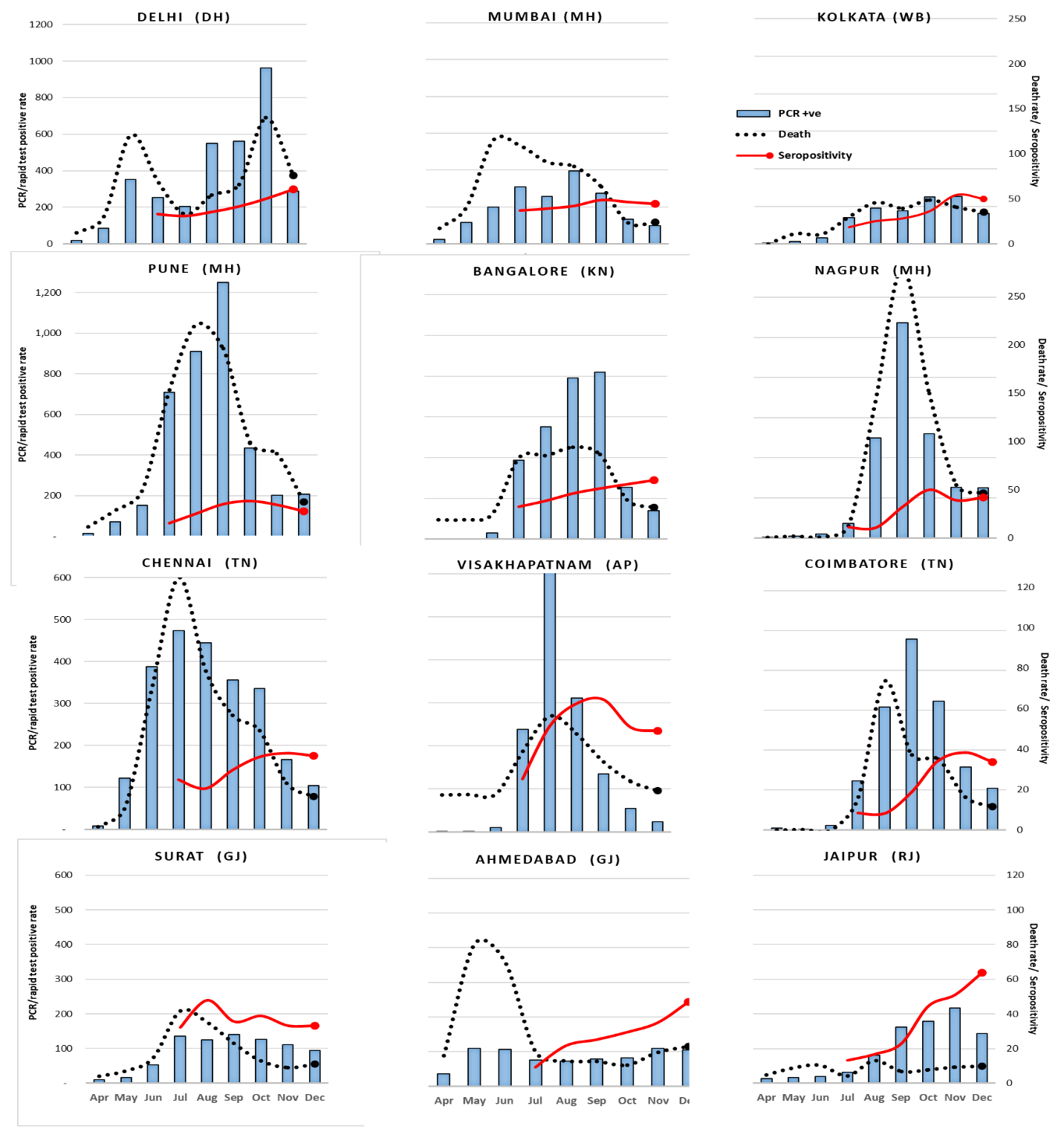

**Supplementary Figure 1 – Variation of monthly assessed PCR/Rapid antigen test positive rates, seropositivity (%) and COVID death rates in 18 metropolitan cities from April-December 2020**

Left-sided axis is the PCR/Rapid test positive rate per 100,000 population. Right-sided axis is for self-referred population IgG seropositivity in percent as well as for COVID death rate per 10,000 population. Mega cities and other metropolitan cites organized separately in order of average PCR/Rapid test positive rate per 100,000 population for April to December. Hyderabad deaths were not available. Pune, Mumbai, Delhi, Kolkata, Bangalore and Nagpur were the highest pandemic affected cities and show a different scale.

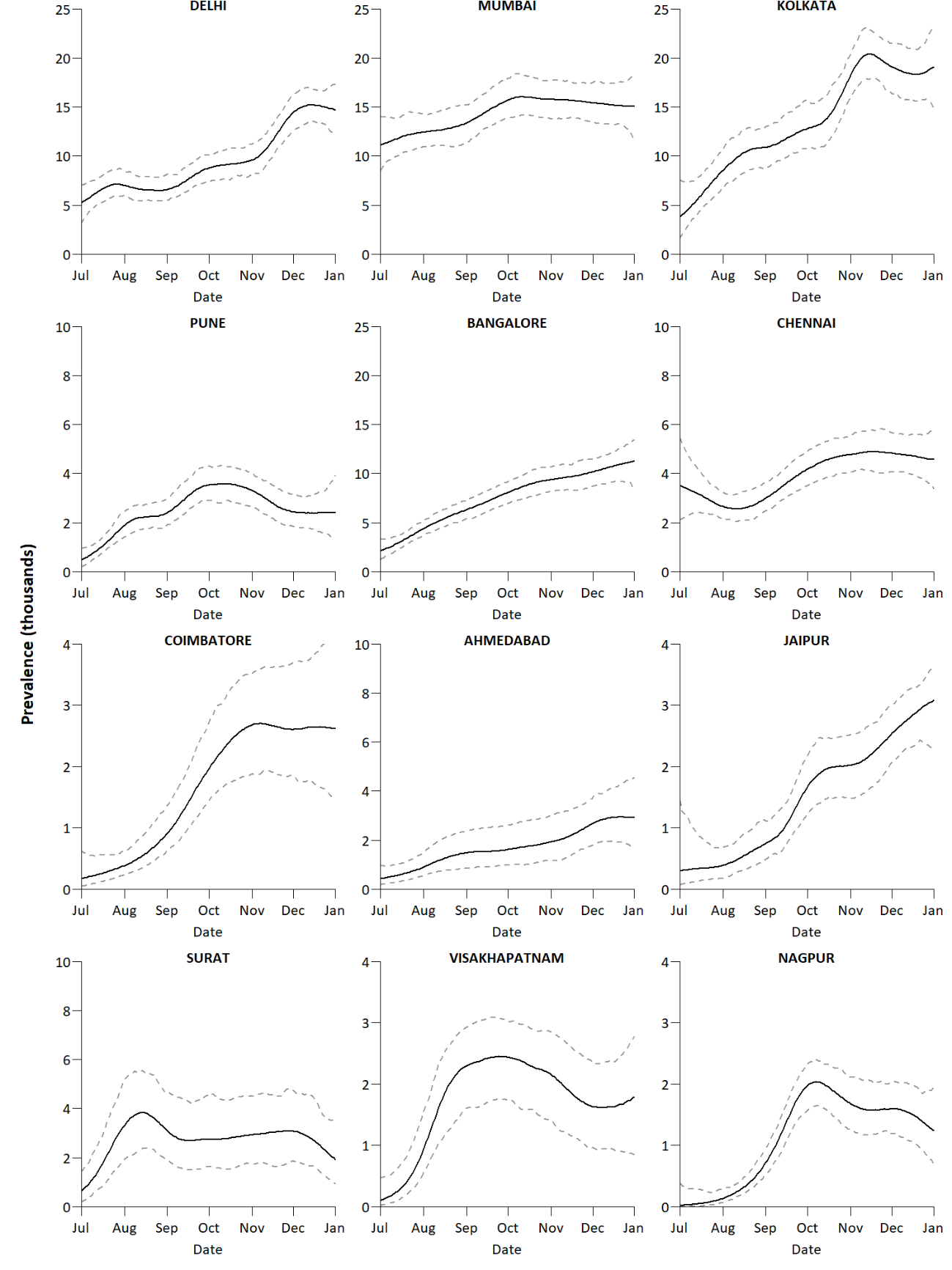

**Supplementary Figure 2 – Seropositivity prevalence for 12 cities from July to December, 2020.**

Seropositivity prevalence were calculated based on a non-spatial logistic regression model, with second order random walks to model the non-linear temporal effects. The logistic regression model estimates were converted to absolute seropositivity prevalence using the total number of serology tests (Supplementary Appendix p4).

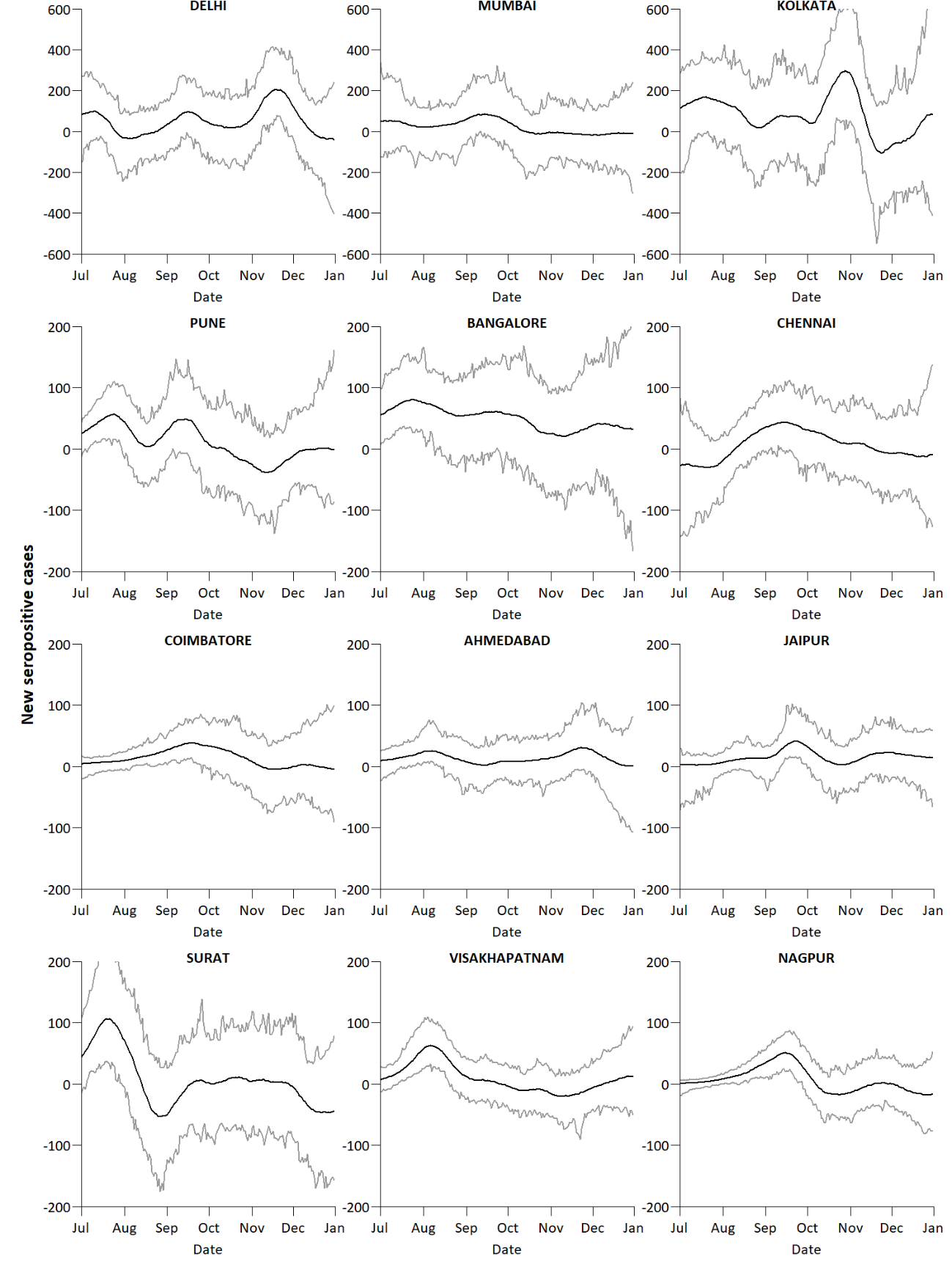

**Supplementary Figure 3 – Daily new seropositive cases for 12 cities from July to December, 2020.**

Daily new seropositive cases were obtained by taking the difference of seropositivity prevalence between consecutive days (Web Figure 3).

**A.**

**B.**

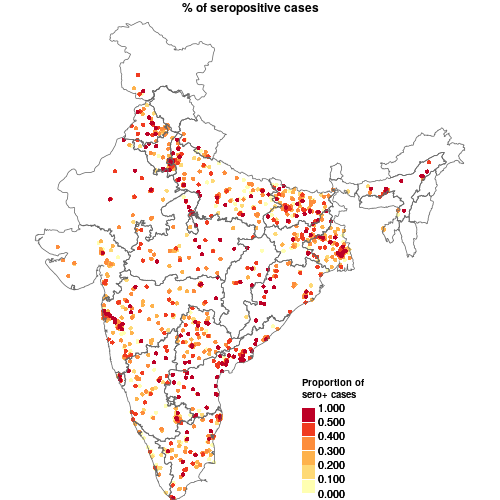

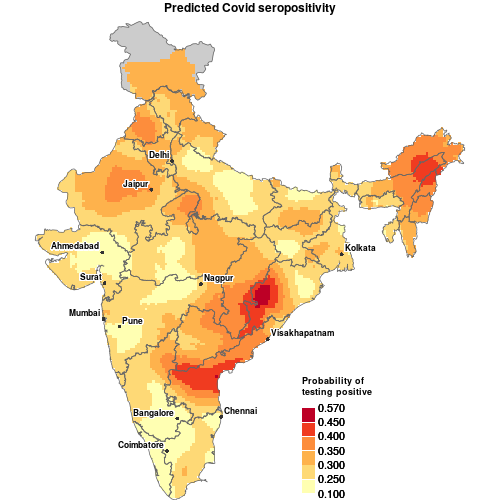

**Supplementary Figure 4 – Distribution of pincodes for serology testing and spatial prediction map based on model-based geostatistical regression analysis of overall seropositivity.**

Panel A: map of % of seropositive cases; Panel B: predicted seropositivity.

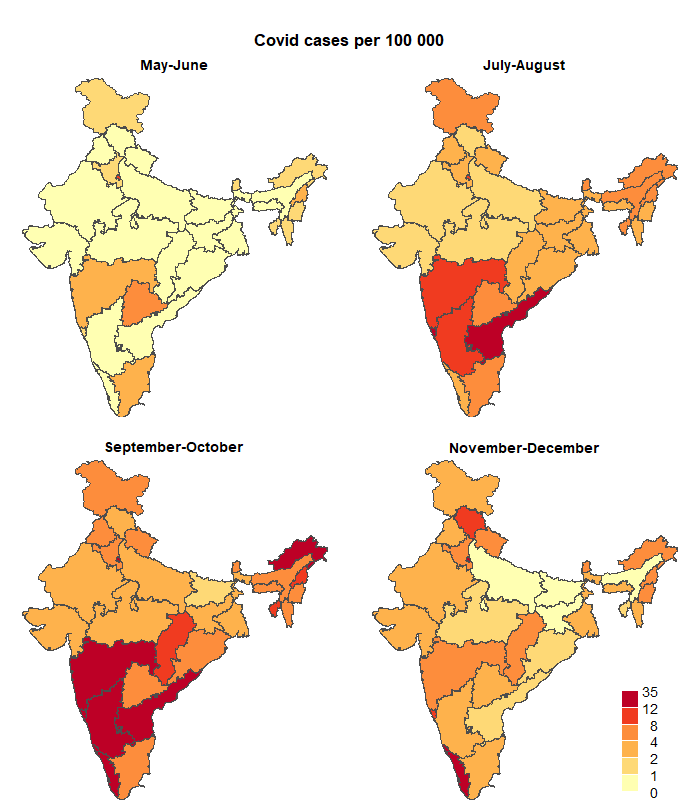

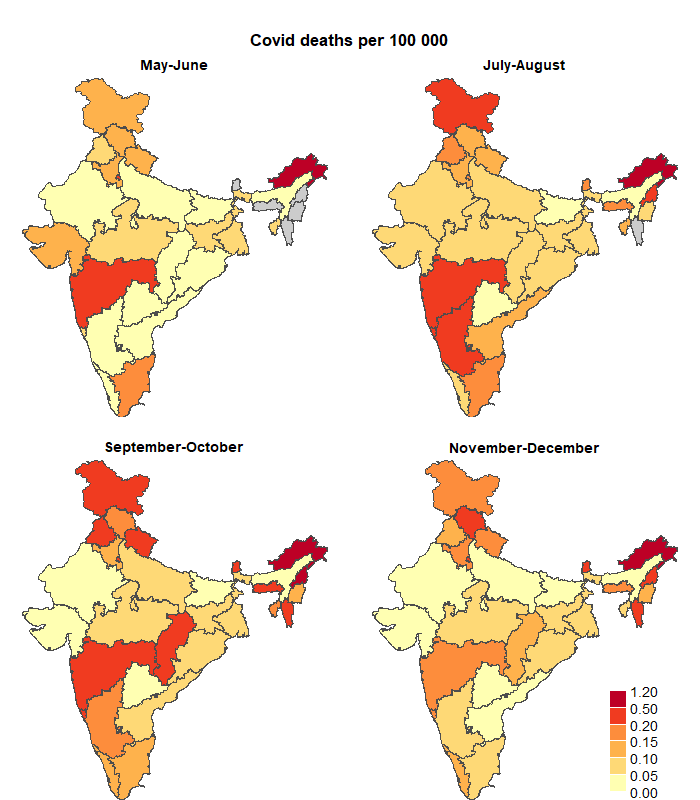

**Supplementary Figure 5 – State-level daily COVID PCR/Rapid test positive cases and deaths per 100 000 by four time periods (May-June, July-August, September-October, November-December)**

Grey shading: data are not available. The daily rates were obtained from a generalized linear model (GLM) with Poisson distribution for the daily COVID cases/deaths. The GLM models had a population offset term and an interaction term between states and time periods, and without the intercept term. The exponentiate of the coefficients represent the daily case/death rates for the combination of states and time periods.

| Current study Seroprevalence | Other study seroprevalence |
| --- | --- |
| 17.1198% | 18.4767% |
| 18.45% | 4.14847% |
| 28.4634% | 39.228% |
|  | 54.1% |
|  | 16.1% |
| 13.3333% | 3.6% |
| 25.9818% | 19.8113% |
| 22.2525% | 19.4872% |
|  | 16.5179% |
|  | 23.494% |
| 21.6859% | 51.5% |
| 32.84% | 6.6% |
| 32.84% | 16.9% |
| 32.84% | 9% |
| 32.84% | 5.2% |
| 28.12% | 27.3% |
| 28.12% | 29.8% |
| 21.6859% | 51.3% |
| 33.4233% | 54.6% |
| 22.84% | 28.39% |
| 27.7606% | 24.08% |
| 34.3263% | 24.11% |
| 50.3% | 56.13% |
| 0.2035% | 50% |
| 0.2035% | 44.8% |
| 0.2035% | 53.8% |

| Current study Seroprevalence | Mean 18 cities | 12 cities 95% LL | 12 cities 95%UL |
| --- | --- | --- | --- |
| 0.1505 | 15.05% | 2.91683% | 27.1833% |
| 0.178043 | 17.8043% | 13.1313% | 22.4774% |
| 0.238243 | 23.8243% | 17.6246% | 30.0239% |
| 0.303796 | 30.3796% | 26.063% | 34.6963% |
| 0.386865 | 38.6865% | 34.9133% | 42.4597% |
| 0.397066 | 39.7066% | 35.7999% | 43.6134% |
| 0.41406 | 41.406% | 35.1039% | 47.7082% |

**Supplementary Figure 6 – Seroprevalence in self-referred populations versus other studies**

Other studies were matched to the setting and month in the self-referred population. The overall prevalence in the self-referred population for the 12 cities is shown in the orange line, with the dotted curves representing 95% confidence intervals based on seropositivity variance of 12 cities at each month.

**References**

1. Babu GR, Sundaresan R, Athreya S, et al. 2020. The burden of active infection and anti-SARS-CoV-2 IgG antibodies in the general population: Results from a statewide survey in Karnataka, India. medRxiv 2020. doi: <https://doi.org/10.1101/2020.12.04.20243949>.
2. Bhatt S, Weiss D, Cameron E, et al. 2015. The effect of malaria control on Plasmodium falciparum in Africa between 2000 and 2015. Nature 526: 207.
3. Brown PE. 2015. Model-based geostatistics the easy way. Journal of Statistical Software 63: 1–24.
4. Brown PE. 2016. Geostatistics in small-area health applications. In Lawson AB, Banerjee S, Haining RP, and Ugarte MD (Eds.), Handbook of spatial epidemiology (pp. 211–224). Chapman; Hall/CRC.
5. Diggle PJ, and Ribeiro PJ. 2006. Model-based geostatistics. New York: Springer-Verlag.
6. George CE, Inbaraj LR, Chandrasingh S, de Witte LP. 2021. High seroprevalence of COVID-19 infection in a large slum in South India; what does it tell us about managing a pandemic and beyond? Epidemiology and Infection 149, e39, 1–6. doi: <https://doi.org/10.1017/S0950268821000273>
7. Ghose A, Bhattacharya S, Karthikeyan AS, et. al. 2020. Community prevalence of antibodies to SARS-CoV-2 and correlates of protective immunity in an Indian metropolitan city. medRxiv 2020. doi: <https://doi.org/10.1101/2020.11.17.20228155>
8. Hammer MS, van Donkelaar A, Li C, et al. 2020. Global Estimates and Long-Term Trends of Fine Particulate Matter Concentrations (1998-2018). Environmental Science & Technology. 2020 Jun 3. doi: <https://pubs.acs.org/doi/10.1021/acs.est.0c01764>
9. Khan SM, Qurieshi MA, Haq I, et al. 2020. Seroprevalence of SARS-CoV-2 specific IgG antibodies in District Srinagar, northern India – a cross-sectional study. bioRxiv 2020. doi: 2020.09.04.282640.
10. Kshatri JS, Bhattacharya D, Kanungo S, et al. 2020. Findings from serological surveys (in August 2020) to assess the exposure of adult population to SARS Cov-2 infection in three cities of Odisha, India. medRxiv 2020. doi: 2020.10.11.20210807
11. International Institute for Population Sciences (IIPS) and ICF. 2017. National Family Health Survey (NFHS-4), 2015-16: India. Mumbai: IIPS.
12. Lindgren F, Rue H, and Lindstrom J. 2011. An explicit link between Gaussian fields and Gaussian Markov random fields: The stochastic partial differential equation approach. J of the Royal Stats Soc B 73: 423–498.
13. Mahapatra P. 2010. An overview of the sample registration system in India. Prince Mahidol Award Conference & Global Health Information Forum. Retrieved from <https://unstats.un.org/unsd/vitalstatkb/KnowledgebaseArticle50447.aspx>
14. Malani A, Shah D, Kang G, et al. Seroprevalence of SARS-CoV-2 in slums versus non-slums in Mumbai, India. The Lancet Global Health. 2021 Feb 1;9(2):e110-1. doi: <https://doi.org/10.1016/S2214-109X(20)30467-8>
15. Mohanan M, Malani A, Krishnan K, et al. 2021. Prevalence of SARS-CoV-2 in Karnataka, India. JAMA. 2021;325(10):1001-1003. doi: 10.1001/jama.2021.0332
16. Murhekar MV, Bhatnagar T, Selvaraju S, et al. 2020. SARS-CoV-2 antibody seroprevalence in India, August–September, 2020: findings from the second nationwide household serosurvey. The Lancet Global Health. 2021 Mar 1;9(3):e257-66. doi: <https://doi.org/10.1016/S2214-109X(20)30544-1>
17. R Core Team. 2018. R: A language and environment for statistical computing. Vienna, Austria: R Foundation for Statistical Computing. Retrieved from <https://www.R-project.org/>
18. Ray A, Singh K, Chattopadhyay S, et al. 2020. Seroprevalence of anti-SARS-CoV-2 IgG antibodies in hospitalized patients at a tertiary referral center in North India. medRxiv 2020. doi: 2020.08.22.20179937.
19. Rue H, Martino S, and Chopin N. 2009. Approximate Bayesian inference for latent Gaussian models by using integrated nested Laplace approximations. Journal of the Royal Statistical Society Series B Statistical Methodology 71: 319–392.
20. Satpati P, Sarangi SS, Gantait K, et al. 2020. Sero-surveillance (IgG) of SARS-CoV-2 among Asymptomatic General population of Paschim Medinipur District, West Bengal, India. medRxiv 2020. doi: 2020.09.12.20193219
21. Sharma N, Sharma P, Basu S, et al. The seroprevalence and trends of SARS-CoV-2 in Delhi, India: A repeated population-based seroepidemiological study. medRxiv. 2020 Jan 1. doi: <https://doi.org/10.1101/2020.12.13.20248123>
22. Siddiqui S, Naushin S, Pradhan S, et al. 2020. SARS-CoV-2 antibody seroprevalence and stability in a tertiary care hospital-setting. medRxiv 2020. doi: 2020.09.02.20186486.
23. Simpson D, Rue H, Riebler A, Martins TG, Sorbye SH. 2017. Penalising model component complexity: A principled, practical approach to constructing priors. Statistical Science 32: 1–28.
24. WorldPop and Center for International Earth Science Information Network, Columbia University. 2018. Global High Resolution Population Denominators Project. doi: <https://dx.doi.org/10.5258/SOTON/WP00649>
